# Emotional Distress, Stress, Anxiety and the Impact of the COVID-19 Pandemic on Early Career Women in Healthcare Sciences Research

**DOI:** 10.1101/2021.12.09.21267476

**Authors:** Noor Bittar, Andrea A. Cohee, April Savoy, Heba M Ismail

## Abstract

**Objectives:** The main objective of this study was to report stress and anxiety levels during the early period of the pandemic on early career women researchers in health sciences research and determine the factors associated with increased stress and anxiety.

**Methods:** A 50-item self-administered internet questionnaire was developed using a mix of Likert-type scales and open-ended response questions. The survey was distributed via email and social media platforms June 10-August 3, 2020. Anxiety and stress associated with the demands of being in health sciences research as well as personal/family demands were assessed through validated measures (Patient Reported Outcomes Measurement Information System (PROMIS)-Anxiety Short Form and Perceived Stress Scale (PSS)) and open-ended responses.

**Results:** One hundred and fifty-one early career women in healthcare sciences research completed the survey. The mean respondent age was 37.3±5.2 years, all had a college degree or higher, 50.3% holding a PhD and 35.8%, M.D. Of the 151 respondents, 128 reported their race/ethnicity and the majority were Caucasian (74.0%). One-third (31.2%) reported being ‘very much’ concerned about reaching their research productivity goals, and 30.1% were ‘very much’ concerned about academic promotion and tenure. Fifty percent reported a ‘moderate’ PROMIS anxiety score and 72.1% reported a ‘moderate’ PSS score. For the open-ended responses, 65.6% reported a worry about their professional goals as a result of the COVID-19 pandemic. Major concerns revolved around finances, childcare and job security.

**Conclusions:** Throughout the pandemic, early career women investigators have reported high overall stress, anxiety and worries.

## Introduction

Early-career women in academia and healthcare sciences research often face multiple stressors and challenges while balancing their personal and professional roles [1]. It is well documented that women in academia struggle with male-dominated institutional cultures, competing family responsibilities, and biases in recruitment, research support, and publication [2]. Unfortunately, with the COVID-19 pandemic, these challenges have been amplified. In particular, those who are engaged in research face delays in their ongoing studies from stay-at-home orders and shutdowns.

Compared to men, many women working from home during the pandemic have reported spending more time on childcare [3]. While most careers have been negatively impacted, early career women in health sciences research have been noted to be at particularly high risk for experiencing a negative impact [4]. Furthermore, those who hold both clinical and academic positions have been either redeployed or asked to perform additional administrative duties, diverting their attention from their academic scholarly work [5, 6]. These challenges have led to a significant decrease in productivity amongst researchers, with 85% of faculty members in one study stating that they expect to decline, postpone or cancel publishing or a research commitment due to COVID-19 [7].

Therefore, it is critical to understand the impact of this pandemic on early-career women in academia to assist in appropriate and targeted support strategies. In this study, we explored the impact of the pandemic on this population. We hypothesized that early-career women health sciences researchers would report greater stress and anxiety related to the pandemic along with increased responsibility and reduced productivity. We aimed to 1) report stress and anxiety levels using validated measures during the early period of the pandemic, and 2) determine factors associated with increased stress and anxiety among these women during the pandemic. We anticipate that these findings will help research institutions in drafting policies to support this population following future pandemics and other natural disasters.

## Methods

### Study Design

This cross-sectional study was submitted, reviewed, and considered an Institutional Review Board (IRB) exempt study by the Indiana University IRB (IRB Protocol #2006034715). A 50-item questionnaire was created through Qualtrics (Qualtrics, Provo, UT) and distributed nationwide via social media platforms, professional organization listservs and emails, and word-of-mouth from June 10th to August 3rd, 2020. No incentives were provided for study participation.

### Eligibility Criteria

A priori inclusion and exclusion criteria were established to identify participants eligible for this study. Inclusion criteria were individuals who identified as women working in health sciences research, at least 15% of their professional time and effort spent on research, able to read and understand English, and in the early stages of their career (research fellows to associate professor level). Those who did not meet any of those inclusion criteria were discontinued from participation in the survey using a gated question method.

### Survey Instrument

The survey instrument (Supplemental materials) combined demographic and career status questions with specific questions regarding the impact of COVID-19 on research, professional support, communication, finances, productivity, childcare and feelings. Additionally, respondents were asked about COVID-19’s impact on their stress and anxiety using validated measures (see *Measures*). Some questions were asked in the form of ‘yes/no’ or ‘select all that apply’ questions, while a majority asked participants to rate their responses on a five-point Likert scale. The questionnaire also included six open-ended questions.

### Measures

#### Demographics

The following demographic data were recorded from survey responses: age, race, ethnicity, degree(s), profession, specialty of practice (if applicable), and average income. Additional data collected included number of years in current position, number of years in research, percent of time spent in clinical, teaching and administrative duties, as well as type of institution (public university, hospital).

#### Stress

Stress was measured using the Perceived Stress Scale (PSS), a 10-question validated measure which asks about the frequency of specific thoughts and feelings in the last month. These included how often respondents felt on top of things, felt able to control irritations in one’s life, felt confident in their ability to handle personal problems, et cetera. Responses were scaled on a 5-point Likert scale (“Never,” “Almost Never,” “Sometimes,” “Fairly Often,” “Very Often”) and were summed to give a result between 0 and 40, with scores of 0-13 indicating low stress, 14-26 moderate stress, and 27-40 high stress [8].

#### Anxiety

Anxiety was measured using the PROMIS Anxiety Short Form, a validated measure of 8 questions evaluating participant feelings over the prior 7 days, specifically assessing how often respondents felt nervous, uneasy, fearful, anxious, et cetera. Questions were asked on a 5-point Likert scale (“Never”, “Rarely”, “Sometimes”, “Often”, “Always”). Responses were summed and scored, converted into T-scores and categorized as “none to slight” (0-55), “mild” (55-59.9), “moderate” (60-69.9) and “severe” (>70) [9].

#### COVID and self-efficacy

Questions also asked about respondents’ confidence in taking care of themselves and household members as well as how much any emotional distress caused by COVID-19 had interfered with self-care and household members’ care. Responses were measured on a 5-point Likert scale (“Not At All”, “A Little”, “Somewhat”, “Quite a Bit”, “Very Much”).

#### COVID and feelings

Eight questions asked about respondents’ feelings secondary to the pandemic. Examples included feeling angry, depressed, anxious, etc. Responses were measured on a 5-point Likert scale (“Not At All”, “A Little”, “Somewhat”, “Quite a Bit”, “Very Much”). Internal agreement between these 8 questions was analyzed using Cronbach’s alpha test (α > 0.8).

#### Open-ended questions and responses

The survey also included open-ended questions. These questions allowed respondents to provide further detail regarding how the COVID-19 pandemic had affected their personal and professional lives. Questions ranged from the most difficult aspects of the pandemic in terms of personal and professional life, worries due to the COVID-19 pandemic to coping mechanisms throughout the pandemic. Individual responses to open-ended questions were quantitatively categorized based on similar responses and grouped according to common themes.

### Data Synthesis

A total of 185 survey responses were collected from this study. One hundred and fifty-one responses met inclusion criteria and 34 were excluded due to being incomplete. All 34 incomplete surveys only answered the first question regarding gender and did not proceed with the remainder of the survey.

Data was summarized for demographic variables including age, race, highest degrees, number of years conducting research, current position, length of time in current position, place of work, and by categories of anxiety and perceived stress. Data were also summarized for variables regarding COVID-19 and research, finances, productivity, diagnosis, and research. The Student’s T-test was used to test for the differences between groups of PROMIS anxiety and PSS stress for continuous variables. Chi-square test was used to test for the differences/correlations for categorical variables. Kruskal-Wallis test was used to determine the correlation between the continuous variables and the categories of anxiety and stress.

Variables that were significant in Chi-square analyses with the PROMIS anxiety and PSS stress scales were then entered into a logistic regression analysis to determine the effect of these variables on prediction of stress and anxiety. All the tests were run at the level of 5% significance. The statistical software SAS 94. (NC, Cary) was used for the statistical analysis of the data.

## Results

### Demographics

One hundred fifty-one women in healthcare sciences research completed the survey. The demographic data for participants is shown in Table 1. The mean respondent age was 37.3±5.2 years. All respondents had a college degree or higher, with 50.3% holding a PhD, 35.8% an M.D, and 11.3% held a combined M.D./PhD. Fifty two percent were classified as Scientists and 47.7% were Physician Scientists. The majority (80.3%) held an Assistant Professor position. At the time of the survey, 72.9% of these women had been in their respective positions for 1-5 years and had been conducting research between 5-10 years (43.1%), with 59.3% noted working at a large public university.

**Table 1.** Demographics and Characteristics.

| **Table 1. Demographics and Characteristics** |  |  |
| --- | --- | --- |
| Mean Age | 37.3±5.2 years |  |
| Race (N=128) |  |  |
| Caucasian | N= 94 | 74.0% |
| Black/AA | N= 5 | 3.9% |
| Hispanic/Latino | N= 6 | 4.7% |
| Asian | N= 14 | 11.0% |
| Multiracial | N= 8 | 6.3% |
| Unspecified | N= 1 | 0.0% |
| Highest Degree (N=151) |  |  |
| MD | N= 54 | 35.8% |
| PhD | N= 76 | 50.3% |
| MD, PhD | N= 17 | 11.3% |
| Masters or less (MPH, MPA, Bachelors) | N= 4 | 2.7% |
| Length of time conducting research (N=151) |  |  |
| < 6 months | N= 3 | 2.0% |
| 1-5 year | N= 50 | 33.1% |
| 5-10 years | N= 65 | 43.2% |
| 15-20 years | N= 28 | 18.5% |
| > 20 years | N= 5 | 3.3% |
| Current Position (N=151) |  |  |
| Associate Professor | N= 16 | 10.6% |
| Assistant Professor/Lecturer/Instructor | N= 99 | 65.6% |
| Post-doctoral/Clinical Fellow | N= 7 | 4.6% |
| Others (Federal Researcher, Admin. Position, Technician, Research Manager) | N= 29 | 19.2% |
| Scientist or Physician (N=151) |  |  |
| Scientist | N= 79 | 52.3% |
| Physician Scientist | N= 72 | 47.7% |
| Time in current position (N=151) |  |  |
| < 6 months | N= 8 | 5.3% |
| 1-5 year | N= 110 | 72.9% |
| 5-10 years | N= 30 | 19.9% |
| 15-20 years | N= 3 | 2.0% |
| > 20 years | N= 0 | 0.0% |
| Place of work (N=151) |  |  |
| Federal | N= 5 | 3.3% |
| Research institute | N= 2 | 1.3% |
| Large public university | N= 89 | 59.3% |
| Large private university | N= 26 | 17.3% |
| Small private university | N= 2 | 1.3% |
| State university | N= 21 | 14.0% |
| Hospital and medical center (academic/non- academic) | N= 5 | 3.3% |

Of the 151 respondents, 128 reported their race/ethnicity, with a majority identifying as Caucasian (74.0%). Out of 132 women who reported their current marital/family status and annual family income, 65.9% were married with children and 44.0% had an annual family income of >$175,000, Table 4.

### Measures of Stress and Anxiety

Results indicated, for overall levels of stress measured using the Cohen’s PSS scale, that 50.0% of participants experienced at least a moderate level of stress, followed by 22.2% with severe levels of stress. Overall levels of anxiety measured using the PROMIS short form questionnaire showcased 72.1% of participants reported moderate levels of anxiety; and 19.4% high anxiety, Table 3. Demographic characteristics were not associated with stress and anxiety in this study.

### COVID-19, Feelings and Baseline Comparison

Participants were asked to rank the pandemic’s impact on their personal well-being through various scaled questions assessing feelings of anxiety, fear, anger, and depression. While there was a varied distribution in responses, a majority stated that there was at least “Somewhat” or “Quite a Bit” of a disruption in their life and mental and emotional well-being as a result of the pandemic. The mean±SD COVID-19 ‘feelings scale’ score was 3.5±0.7 which was highly reliable with a Cronbach’s Alpha of 0.85. Negative feelings towards COVID-19 showed significant positive associations to both PSS and PROMIS Anxiety scores and the degrees of stress and anxiety (p<0.01 for both), with higher scores associated with both moderate and severe anxiety and stress, supplementary table 1.

#### Baseline Stress and Anxiety

When participants were asked to rank their levels of anxiety and stress during the COVID-19 pandemic in comparison to before the pandemic, a majority (58.3%) of participants scored levels of stress and anxiety during the pandemic as “More than Before”, while 26.5% scored “Much more than before”. Similarly, 51.5% reported a stress level “More than Before” and 33.3% reported a stress level “Much more than before” the pandemic.

### Measures of Self-Efficacy

Thirty two percent of respondents felt that COVID-19 interfered “A Little” with their ability to take care of themselves. When adding burdens of social distancing to their self-care, a large percentage (36.9%) stated that they felt “Quite a Bit” confident. Of the 32 women who answered how their COVID-related emotional distress interfered with the management of their household with special needs, 31.3% stated there was “Somewhat” of an interference. Lastly, 53.8% of 26 women answered that they were “Somewhat” confident in managing household members with special needs, Table 2. With the exception of associations of responses to the question “How confident are you that you can take care of yourself with the added burden of social distancing?” and PROMIS anxiety scores (X^2^= 13.9, p=0.31) and the question “How confident do you feel that you can manage your household members with special needs at home?” and the PSS stress scores (X^2^=19.8, p=0.07), all other associations showed a positive trend and were statistically significant (p<0.05), supplemental table 2.

**Table 2:** Participant responses to self-efficacy questions regarding confidence and distress during the COVID-19 pandemic.

| *COVID-19 and Self-Efficacy* | *Not at all* | *A little* | *Somewhat* | *Quite a bit* | *Very much* |
| --- | --- | --- | --- | --- | --- |
| How confident are you that you can take care of yourself with the added burden of social distancing?<br>**N=130** | 2.31%<br>(n=3) | 7.69%<br>(n=10) | 20%<br>(n=26) | 36.9%<br>(n=48) | 33.1%<br>(n=43) |
| How much does emotional distress caused by COVID-19 interfere with taking care of yourself?<br>**N= 130** | 13.1%<br>(n=17) | 32.3%<br>(n=42) | 30.8%<br>(n=40) | 16.9%<br>(n=22) | 6.9%<br>(n=9) |
| How much does emotional distress caused by COVID-19 interfere with the management of household members with special needs?<br>**N= 32** | 15.6%<br>(n=5) | 15.6%<br>(n=5) | 31.3%<br>(n=10) | 25.0%<br>(n=8) | 12.5%<br>(n=4) |
| How confident do you feel that you can manage your household members with special needs at home?<br>**N=26** | 3.9%<br>(n=1) | 3.85%<br>(n=1) | 53.9%<br>(n=14) | 26.9%<br>(n=7) | 11.5%<br>(n=3) |

**Table 3:** Responses to Anxiety and Stress Scales:

|  | **Number** | **Frequency** |
| --- | --- | --- |
| **PROMIS anxiety scale** |  |  |
| None to slight | 20 | 15.9 |
| Mild | 15 | 11.9 |
| Moderate | 63 | 50.0 |
| Severe | 28 | 22.2 |
| **PSS stress scale** |  |  |
| Low | 11 | 8.5 |
| Moderate | 93 | 72.1 |
| High | 25 | 19.4 |

### Impact on Research and Institutional Support

While many participants who stated lab closures or research projects were on hold due to the COVID-19 pandemic, there were no significant correlations to stress and anxiety levels among participants. The same was true for participants who stated that they had taken on more responsibilities due to the pandemic. However, for those who stated the highest average amount of clinical effort, there were negative associations with anxiety levels as measured by PROMIS scores (p = 0.05), but no associations with stress levels. When reporting COVID-19-related concerns with research goals and productivity, 28.9% reported that they were ‘quite a bit’ concerned and 45.9% were ‘very much’ concerned.

A majority of participants stated that their productivity decreased with either increased or decreased work hours. Of those who reported ‘very much’ concerned, 29.0% (n=18) had severe anxiety and 27.4% (n=17) had high stress levels. Of the participants who stated their productivity decreased while their work hours were reduced, the majority had moderate to high anxiety and stress levels, comprising 64.0% of those in the High Stress category and 64.3% in the Severe Anxiety category, supplemental table 3. Additional concerns of promotion/tenure, current and future funding, or salary cuts and furloughs were stated; however, there were no significant differences among the groups. Additionally, 22.0% reported no-to-little support from their institution, while the majority (34.1%) felt somewhat supported by their institution. Conversely, 35.1% felt supported by their department within the institution.

### Impact on Finances and Responsibilities

For COVID-19 and finances questions, there was a significant difference in stress levels by total family income (p=0.02), but not in anxiety levels. The majority (47.3%) of those in the moderate stress level as measured by PSS were in the mid-range income level of 99,000-175,000. Interestingly, while many mentioned childcare obligations as factors associated with their stress and anxiety levels in the open-ended questions, there were no significant differences among participants’ levels of stress and anxiety when reporting school-aged children in their household, responsibility level and/or being in charge of the e-learning, Table 4. No significant association with stress and anxiety scores appeared with other COVID-19 and financial questions within the survey.

**Table 4:** COVID-19, Finances and Responsibilities’ Responses.

| **Questions** | **Number** | **Frequency** |
| --- | --- | --- |
| **Have there been any salary cuts or furloughs to you, your staff, or anyone in your research team?** |  |  |
| Yes | 21 | 15.9 |
| No | 111 | 84.1 |
| **Have you been asked to redeploy to clinical duties or provide information on willingness to redeploy** |  |  |
| Yes | 51 | 38.6 |
| No | 41 | 31.1 |
| N/A | 40 | 30.3 |
| **Have you experienced financial difficulty related to COVID-19** |  |  |
| Yes | 18 | 13.6 |
| No | 114 | 86.4 |
| **Total annual family income** |  |  |
| <99,000 | 17 | 12.9 |
| 99,000-175,000 | 56 | 42.4 |
| >175,000 | 59 | 44.7 |
| Missing | 19 |  |
| **What is your current marital/family status?** |  |  |
| Married without Children | 23 | 17.4 |
| Married with Children | 87 | 65.9 |
| Single without children | 18 | 13.6 |
| Single with children | 4 | 3.0 |
| Missing | 19 |  |
| **Who lives in your household** |  |  |
| Adults | 39 | 29.6 |
| Adults,Children | 88 | 66.7 |
| Adults,Children,Seniors (> 65 Years Old) | 3 | 2.3 |
| Missing | 21 |  |
| **Children in the household** |  |  |
| Children < 5 Years Old | 32 | 34.8 |
| Children < 5 Years Old,Children Between 5 and 12 Years | 31 | 33.7 |
| Children < 5 Years Old,Children Between 5 and 12 Years,Teenagers < 18 Years Old,Children with Special Needs | 3 | 3.3 |
| Children Between 5 and 12 Years and teenagers | 26 | 28.3 |
| Missing | 59 |  |
| **Are/were children receiving e-learning at home during the Pandemic** |  |  |
| Yes | 68 | 73.9 |
| No | 24 | 26.1 |
| **Are/were you in charge of schooling and e-learning?** |  |  |
| Yes, full time | 27 | 39.7 |
| Yes, part time | 2 | 36 |
| No | 3 | 5 |
| **Due to the effects of the COVID-19 Pandemic, the responsibility of childcare (or adult/senior care) has now** |  |  |
| Fallen mostly on me | 28 | 30.1 |
| Is currently equally shared with others | 32 | 34.4 |
| Is currently unequally share with others (others do most of it) | 6 | 6.5 |
| Is currently unequally shared with others (I do most of it) | 27 | 29.0 |

### Impact of COVID-19 diagnosis

Regarding whether participants or someone they knew had been diagnosed with COVID-19, the majority (54.5%, n=72) said “No” with 86.4% of them reporting not currently caring for any patients with COVID-19 in their practice.

Interestingly, those who responded yes to each of the questions related to ‘knowing or having COVID-19’ or ‘caring for someone diagnosed’ in their practice also showed moderate to high levels of anxiety with a significant positive association with PROMIS Anxiety scores (p=0.03 and p=0.02, respectively). There were no associations seen with stress levels and there were no associations seen between active or passive COVID-19-related information seeking behaviors and anxiety or stress levels.

### Regression Models

With the reference group being the ‘severe’ PROMIS anxiety scores, the odds of having low anxiety scores were greater if the participant felt supported by their institution (none to low anxiety scores OR=2.2, CI 1.19-4.08, p=0.01) or by their department (none to low anxiety scores OR=2.29, CI 1.23-4.26, p=0.01; mild anxiety scores OR=1.91, CI 1.01-3.62, p=0.04). Whereas the odds of increased time and percent effort spent on administrative tasks was lower in the ‘moderate’ stress level group compared to the ‘high’ stress level group as measured by PSS, (OR=0.95, CI 0.91-0.99, p-value 0.02).

### Additional open-ended questions

#### Career impacts

A large portion of the sample identified the effect on their career (n=74/131; 56.5%), including reduced research work hours and productivity (n=83; 63.4%). Reasons for reduced productivity included lack of focus and distraction (21.1%), stress (17.5%), and work hours being reduced (14.7%).

#### Worries

The majority (65.6%) of women reported a worry about their professional goals as a result of the COVID-19 pandemic. Major concerns revolved around finances, childcare and job security. Financial status was addressed either on a personal level within their household or professional level due to grant cuts and grant deadlines not being met. Sixty-eight percent reported their biggest professional concerns, with maintaining productivity and career impacts at the top, supplemental table 4.

#### Challenges

A majority of respondents (n=97, 64.2%) shared the most challenging aspects of managing their personal and professional lives during the COVID-19 pandemic, with additional responsibilities (childcare, added clinical hours) encompassing 29.0% of responses.

Childcare was a major theme among participants. In fact, 131 stated that they had children under 18 years of age in their households. Of those with children, 47.8% were younger than 5 years old and 41.0% were between 5-12 years of age. Eighty three percent (N=68/92) of children were receiving e-learning and 39.7% of participants reported that they oversaw full-time learning (n=27).

#### Coping Mechanisms

We also inquired about participants’ coping mechanisms for stress and anxiety. The majority (n=103, 68.2%) responded to the question on whether they have someone to talk to about their concerns with 91 (88.3%) of women saying that they do have someone to speak with in some capacity. This encompassed 73.6% mentioning friends (colleagues, friends, peers), 38.5% (n=35) a mentor or supervisor, and 15.4% (n=14) mentioning family members (family, relatives, spouse). Other outlets included social media and online platforms (4.4%), therapists (3.3%) or an unspecified source (2.2%). A small number (n=13) of respondents mentioned having someone to speak to about their concerns but not being able to reach out to them due to lack of time or believing they would not understand their concerns. Further, 11 other respondents said they did not have someone to voice concerns to.

The majority of respondents (n=96, 63.6 %) reported on how they were coping with the stress associated with the pandemic. Coping mechanisms included physical activity (55 respondents), spending time with and talking with family and friends (41 respondents), optimism and leisure activities (n=22), keeping up with a routine (n=18) and eating and drinking alcohol (n=20). Other less frequent coping strategies included limited social media use and therapeutic interventions such as meditation or therapy (n=3). A small number (n=5) of respondents stated they were not coping well or were overwhelmed with worry and childcare.

## Discussion

This study aimed to provide insight into the impact of the COVID-19 pandemic on women in academia in an effort to provide more equitable institutional policies in the field of science. Our results show that the majority had moderate anxiety and stress levels. About a third were ‘very much’ concerned about reaching their research productivity goals, academic promotion and tenure. About two thirds reported a worry about their professional goals as a result of the COVID-19 pandemic and major concerns revolved around finances, childcare and job security.

Here, we focused specifically on the increased level of stress and anxiety that academic women have encountered as a result of their professional obligations becoming further intertwined with family and other personal responsibilities. These problems had amplified due to budget cuts, furloughs, redeployment, additional responsibilities and concerns for long-term career impacts, consistent with what others have reported [4]. Academic women are challenged regularly with daily stressors, without the addition of a pandemic’s implications on their personal and professional well-being. Our predictions were supported in that women in academia reported higher levels of stress and anxiety as a result of the pandemic compared to prior to the pandemic.

This work highlights the issues women in academic research settings encountered during the pandemic. Compared to before the pandemic, women have reported increased overall stress due to their household setting, additional responsibilities, and financial concerns. Children were in school while parents would be at work, daycare and babysitting services were available and affordable, and afterschool routines were manageable prior to the pandemic. Further, researchers have had to allocate their work hours towards more clinical or administrative work rather than their ongoing projects, and grant money and proposal deadlines have become difficult to reach. In addition, prior to the pandemic, grant proposals were underway and fairly distributed, but as seen in our results, this has become a major concern for many women in academics, contributing therefore to higher stress and anxiety levels [5, 7]. These effects may hinder any long-term progression of women’s trajectory in academic medical research.

We can learn from previous pandemics and recession where the impacts on women in academic resulted in pay gaps increasing by 3.7% for women in addition to a decline in leadership roles as women are overlooked due to time constraints and redirection to outside obligations during times of crises [10, 11]. Our data illustrates a need for attentive care of academic working women, who feel under-supported, unheard and mispresented in their respective field of work, especially when compared to their male counterparts or colleagues without children [12-14] and/or additional obligations outside of work. Academic institutions and funding organizations should consider providing strategic support, equitable extensions and allocated funding to those unfairly impacted by crises. Only then can there be equal opportunity for those in academic appointments. This pandemic has shed a light on the policies that may need to be put in place to better prepare and support women in academia, whether it be extensions on grants deadlines, increased administrative support and alternatives to delayed research with added clinical responsibilities or administrative tasks that have needed to be taken on by these women [10, 11].

It is also important to address the outliers of this study that were brought to light as a result of the COVID-19 pandemic. A few women voiced concerns in regard to issues on social and racial injustices (data not shown) coinciding with the onset of the pandemic. Institutions should want to look into providing those who feel underrepresented and supported in the workplace with an uplifting support network. As participants mentioned these issues to be affecting their stress levels on both a professional and personal level, institutions should be aware of the emotional impacts social and racial injustice have on their faculty and implement effective means of support; be it by providing outlets for open discussion and a communitive environment to colleagues with contacts and resources to reach out to if even in need.

Women who feel misunderstood by their colleagues, either because they do not have children, are males or can simply not relate to their situation, also need to be recognized. Women in our study are stating that they are uncomfortable or afraid to speak up about their concerns. Institutions may want to address these issues by implementing social support systems and routine supervisor-colleague sessions. Women should have a place where they are able to bring about concerns without judgement or fear of potential career impacts. Trainings should be implemented in the workplace to allow for unbiased, active discussions on how to best address the concerns of women in academic appointments. One suggestion would be to implement a peer group within the workplace so that women can connect with those within the institution that they can most relate to, and if it is not available, there should be administrative efforts made to support these women and allow them to address concerns openly and freely.

Overall, it is critical to take into consideration the impact of the COVID-19 pandemic on women in academia. By opening up the conversation on these topics of concern, effective change in the workplace on both an institutional and systematic level can be made. Allocation of and access to resources for those struggling with abrupt work-life alterations should be made available at all times. This would help prevent these concerns from arising and interfering with the work-life balance for so many individuals struggling to remain in academia and research. It is important to have support networks and flexible policies in place to protect those in academic appointments from unhealthy stress and potential career insecurity, especially for future crises that may yet again, unexpectedly arise. Redefining measures of scholarly productivity and providing flexible policies should be implemented for these special groups.

This study is not without limitations. This survey was conducted early during the pandemic but after the shutdown mandate was removed in most states. Thus, the sentiments and stressors may have changed over time. This study also assessed marital status with childcare and clinical responsibility allocations. While there was a majority correlation, it should be noted that our respondent population was majority married with children, thus impacting the generalizability of our results. The data collected also consisted of majority Caucasian women, so differences across ethnic groups and perceptions cannot be generalized, and significant correlations could not be made. In addition, as only females were surveyed, adequate correlations to men’s perceptions of the COVID-19 impact were not collected. While many women stated their concerns of being compared to their colleagues or unfair advantages, we cannot say for certain that men do not also hold similar concerns, or that it solely applies to women. Men may also have financial concerns, time management and family obligations or childcare responsibilities that were not noted in this study. In addition, we could not identify a normative comparator group to compare levels of stress and anxiety to. This is because every field is highly specialized and results from one group may not be applicable to others. Yet, it is still important to emphasize the pandemic concerns mentioned in this study to better establish policies and support networks that would benefit and create a level playing field for all individuals involved. This study also measured several aspects and potential variables contributing to stress and anxiety and surveyed a good representative number of women in academia engaged in health sciences research across the country.

This study focused on early career women in academia. It would be interesting to look further into those in later academic standings, to see if their opinions on the pandemic--whether their standing in respective research appointments and their perception of the impact on their careers—are comparable to the majority assessed in this study. As this study did not survey men, it would be a valuable asset to distribute and compare their opinions and perceptions of COVID-19 on their career journeys as well. Lastly, by expanding this survey over a longer time period to more age groups, ethnicities and marital statuses, a more diverse population would be beneficial in assessing whether or not there were differences across groups. These future modifications would add to the generalizability and significance of our study correlations and bring about further necessary conversations.

Overall, this study indicated there were greater levels of stress, worry and challenges of work-life balance compared to before pandemic. Avenues for support of the work-life dynamic has been uprooted by pandemics and academic institutions and funding organizations should consider providing strategic support, equitable extensions and allocated funding to academic women.

## Data Availability

All data produced in the present study are available upon reasonable request to the authors

## Acknowledgements

This publication was also made possible with support from Grant Numbers, KL2TR002530 (B. Tucker Edmonds, PI), and UL1TR002529 (S. Wiehe and S. Moe, co-PIs) from the National Institutes of Health, National Center for Advancing Translational Sciences, Clinical and Translational Sciences Award.

## Data Availability

Data from this study is available upon request from the authors.

## Contribution statement

HMI conceptualized the study. NB, HMI, AC and AS analyzed and interpreted the data, and wrote the manuscript. All authors contributed to the design, interpreted the data and reviewed/edited the manuscript. HMI is the guarantor for this study.

